# Mapping First to Second wave transition of covid19 Indian data via Sigmoid function and prediction of Third wave

**DOI:** 10.1101/2021.07.11.21260325

**Authors:** Supriya Mondal, Sabyasachi Ghosh

## Abstract

Understanding first and second wave of covid19 Indian data along with its few selective states, we have realized a transition between two Sigmoid pattern with twice larger growth parameter and maximum values of cumulative data. As a result of those transition, time duration of second wave shrink to half of that first wave with four times larger peak values. It is really interesting that the facts can be easily understood by simple algebraic expressions of Sigmoid function. After understanding the crossing zone between first and second wave curves, a third wave Sigmoid pattern is guessed.

## 1 Introduction

Spreading of the novel coronavirus, covid19, from China to entire globe become so alarming that the World Health Organization (WHO) declared it as a pandemic disease on 11th March 2020 [1, 2]. The data of covid infected, recovered and death are maintained by different countries in their government based websites. Ref. [3] is citing the corresponding website for India. From 2020 to now, a huge amount of works are attempted to fit the covid infected daily cases and cumulative data for predicting the their pattern. Few of them are cited in Refs. [4, 5, 6, 7, 8, 9, 10, 11, 12, 13, 14, 15, 16, 17, 18, 19, 20, 21, 22, 23, 24, 25, 26, 27, 28, 29], where few Refs. [18, 24] work on second wave data and third wave prediction. Most of the studies are based on SIR model [30, 31, 32] and its extensions. This framework is widely used for other countries, e.g. Ref. [33]. However, a simple logistic function description like Sigmoid function [34, 35, 36] can also be an easy-dealing tools to understand the epidemic outbreak. In our earlier works [4, 5, 6], we have used this Sigmoid function framework for predicting first wave of India data. This framework is also used for understanding epidemic size of other countries, for example Refs. [35, 36, 37].

Present work is aimed to explain the existing first and second wave covid19 infection data with the help of two different Sigmoid functions and analyzing their trends, we have predicted third wave pattern. The article is organized as follows. Starting with brief formalism part of Sigmoid function, we have discussed about the steps of generating curves in the Sec. (2). Next, in results section, we have described the first and second wave curves including our third wave predicted curves. After analyzing those results, at the end we summarized our work.

## 2 Mathematical Framework

In this framework part, we will discuss quickly about the Sigmoid function which will be used to interpret covid 19 data. Then we will discuss about the steps, through which we proceed.

The form of Sigmoid function is

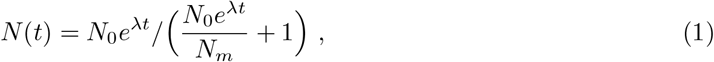

where *N*_0_ is initial number of cases, *λ* is growth parameter, *N*_*m*_ is the maximum values, where cumulative case *N* (*t*) will be saturated. Here t represents number of days. Now, the time derivative of Sigmoid function is

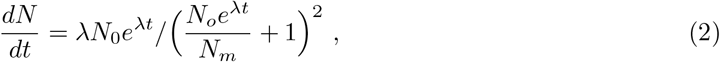

which is the number of new cases per day as we see in covid 19 data. Sigmoid function shows exponential behaviour in low values of *t* but it will saturate to a maximum values (*N*_*max*_) at high values of *t*. When we analyze its derivative or slope, then we will get first increasing and then decreasing trends after showing a peak. The peak structure of daily cases depends on three parameters *N*_*m*_, *N*_0_, *λ*. The peak time *t*_*p*_, when daily cases reach its highest value, can be expressed as

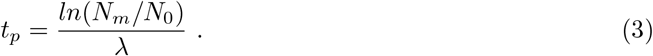

At *t* = *t*_*p*_ daily cases and cumulative cases are respectively

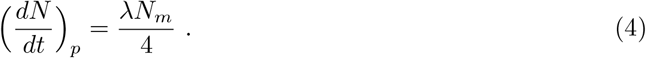

and

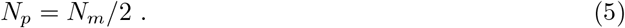

Above simple formalism can be useful to describe covid 19 data pattern. In India we found two waves whose cumulative and daily cases data patterns can expressed in terms of two consecutive Sigmoid functions and their time derivatives.

From the first wave of Covid 19 Indian data [3] we find out the values of *t*_*p*1_, (*dN/dt*)_*p*1_ and *N*_*m*1_. These values are used in Eqs. (3) and (4) to find out the values of *λ*_1_ and *N*_01_. Subscript 1 is added in the notations of different parameters to assign first wave case. For India and some selective states - 1) Maharashtra (MH), 2) Kerala (KL), 3) Karnataka (KA), 4) Tamil Nadu(TN), 5) Andhrapradesh (AP), 6) West Bengal(WB), those parameters are tabulated in Table. (1).

**Table 1:**
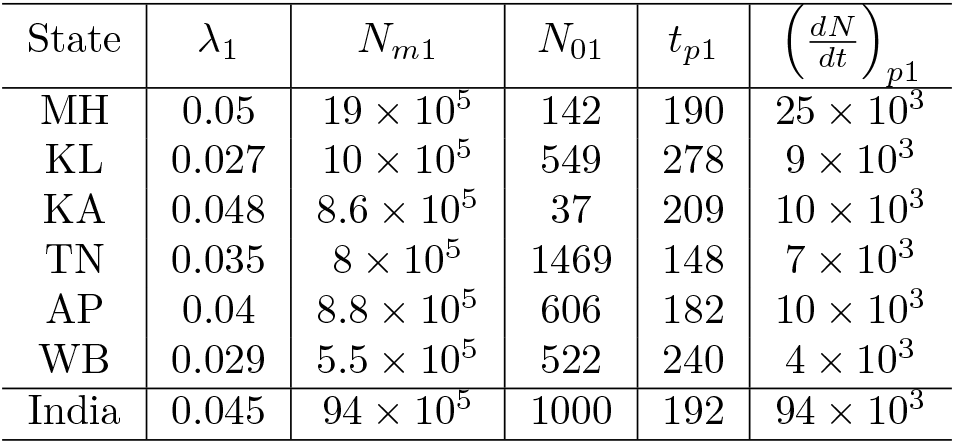
Different parameters of Sigmoid functions, which can grossly describe covid19 first wave data of India and selective states - MH, KL, KA, TN, AP, TN.

| State | $\lambda_1$ | $N_{m1}$ | $N_{01}$ | $t_{p1}$ | $\left(\frac{dN}{dt}\right)_{p1}$ |
| --- | --- | --- | --- | --- | --- |
| MH | 0.05 | $19 \times 10^5$ | 142 | 190 | $25 \times 10^3$ |
| KL | 0.027 | $10 \times 10^5$ | 549 | 278 | $9 \times 10^3$ |
| KA | 0.048 | $8.6 \times 10^5$ | 37 | 209 | $10 \times 10^3$ |
| TN | 0.035 | $8 \times 10^5$ | 1469 | 148 | $7 \times 10^3$ |
| AP | 0.04 | $8.8 \times 10^5$ | 606 | 182 | $10 \times 10^3$ |
| WB | 0.029 | $5.5 \times 10^5$ | 522 | 240 | $4 \times 10^3$ |
| India | 0.045 | $94 \times 10^5$ | 1000 | 192 | $94 \times 10^3$ |

Next, when we will go for corresponding second wave data, we will not get *N*_*m*2_ as it is not till finished and we can not see the second saturated cumulative values. However, we can see the *N*_*p*2_ values from data and by using Eq. (5), we can guess *N*_*m*2_ by making twice of *N*_*p*2_. Here, subscript 2 is added in the notations of different parameters to assign second wave case. Another important point for cumulative data of second wave is that we will redefine it by subtracting first wave maximum values *N*_*m*1_. It means that Eqs. (1) and (2) for second waves will be

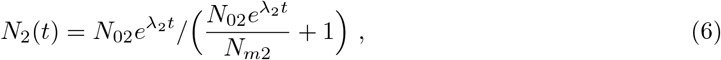

and

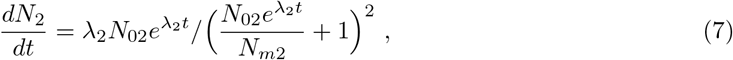

where *N*_*m*2_ = (*N*_*p*2_ −*N*_*m*1_)*×*2. So knowing *N*_*m*1_, *N*_*p*2_ from data, we can guess about *N*_*m*2_. Although, we should keep in mind that *N*_*m*1_ + *N*_*m*2_ is actual saturated values of second wave case, when we compare it with actual data. The parameters of second waves for India and the selective states are given in Table. (2).

**Table 2:**
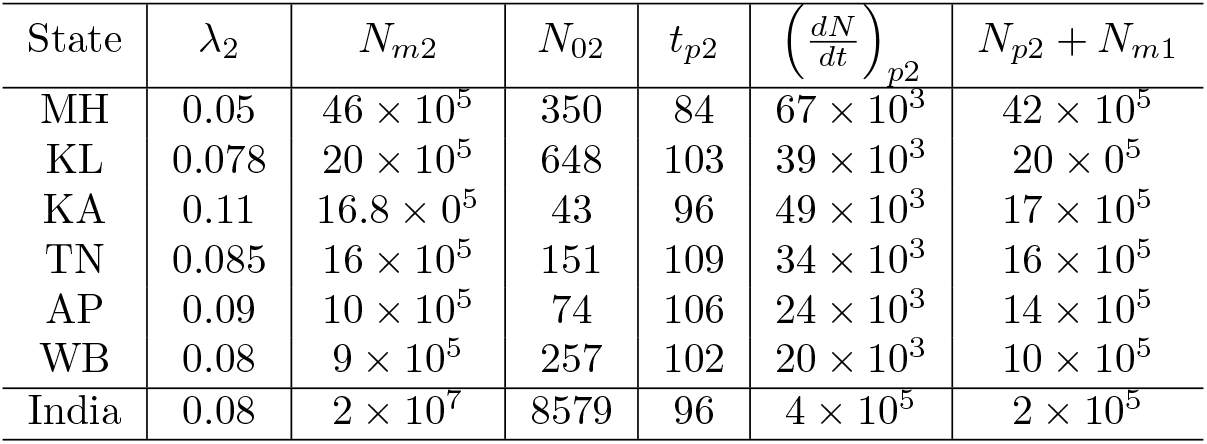
Same as Table (1) for second wave.

| State | $\lambda_2$ | $N_{m2}$ | $N_{02}$ | $t_{p2}$ | $\left(\frac{dN}{dt}\right)_{p2}$ | $N_{p2} + N_{m1}$ |
| --- | --- | --- | --- | --- | --- | --- |
| MH | 0.05 | $46 \times 10^5$ | 350 | 84 | $67 \times 10^3$ | $42 \times 10^5$ |
| KL | 0.078 | $20 \times 10^5$ | 648 | 103 | $39 \times 10^3$ | $20 \times 10^5$ |
| KA | 0.11 | $16.8 \times 10^5$ | 43 | 96 | $49 \times 10^3$ | $17 \times 10^5$ |
| TN | 0.085 | $16 \times 10^5$ | 151 | 109 | $34 \times 10^3$ | $16 \times 10^5$ |
| AP | 0.09 | $10 \times 10^5$ | 74 | 106 | $24 \times 10^3$ | $14 \times 10^5$ |
| WB | 0.08 | $9 \times 10^5$ | 257 | 102 | $20 \times 10^3$ | $10 \times 10^5$ |
| India | 0.08 | $2 \times 10^7$ | 8579 | 96 | $4 \times 10^5$ | $2 \times 10^5$ |

## 3 Results

We have described the steps, through which we have find the parameters of first and second wave of covid 19 spreading. Fig. (1) shows the Sigmoid function nature of cumulative *N* and daily case *dN/dt* data of India. In the left and right panels of Fig. (1) represents the data points (squares and circles) and corresponding Sigmoid fitted curves (dotted and solid lines) in first and second waves respectively. We consider 3 main data points of daily and cumulative cases at *t* = (*t*_*p*_ − 2*/λ*), *t*_*p*_, (*t*_*p*_ +2*/λ*), within which spreading become most dominant. We have taken three different values of *λ* = 0.04, 0.045 and 0.05 to fit the three data points of first wave. In another aspect, Second wave is well fitted with one *λ* = 0.08. First waves is saturated in *N*_*m*1_ = 10^7^ and second waves is saturated in *N*_*m*2_ = 2 × 10^7^ which are already seen in covid 19 data. They are implemented as important inputs to build corresponding Sigmoid curves.

**Figure 1:**
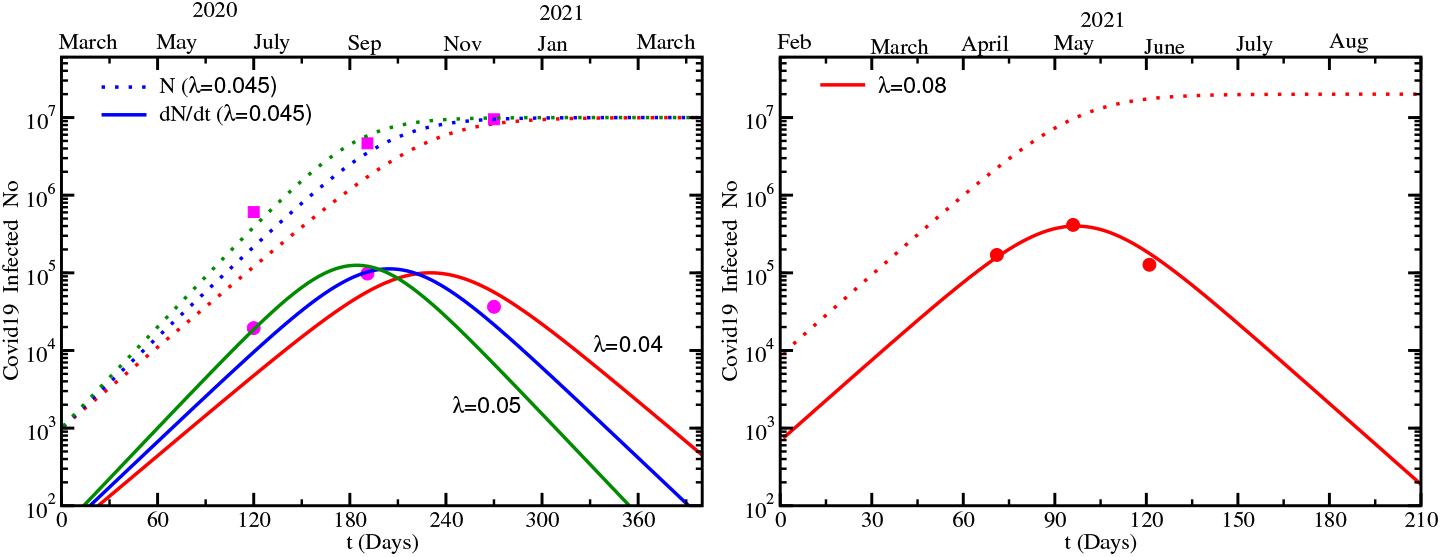
Left panel: Cumulative(squares) and daily cases (circles) data, fitted in Sigmoid curves(dotted line) and their derivatives (solid lines) for first wave. Right panel: Same for second wave.

Next, we will generate similar graphs in Figs. (2) and (3) for 6 selective states - MH, AP, WB, TN, KL and KA.In Fig. (2) we noticed that the daily cases data of MH, AP and TN in first wave are well fitted by (time derivative of) Sigmoid function but the same for WB, KA and KL are not so well fitted by Sigmoid function. On the other hand in second wave all those data are favouring the Sigmoid function, which can be seen in Fig. (3). In first wave there was no sharp peak for few states where as peak was clearly seen during second wave almost in every states. This is most probably because of rapidly growing of daily cases in second wave which was lacking for few states in first wave.

**Figure 2:**
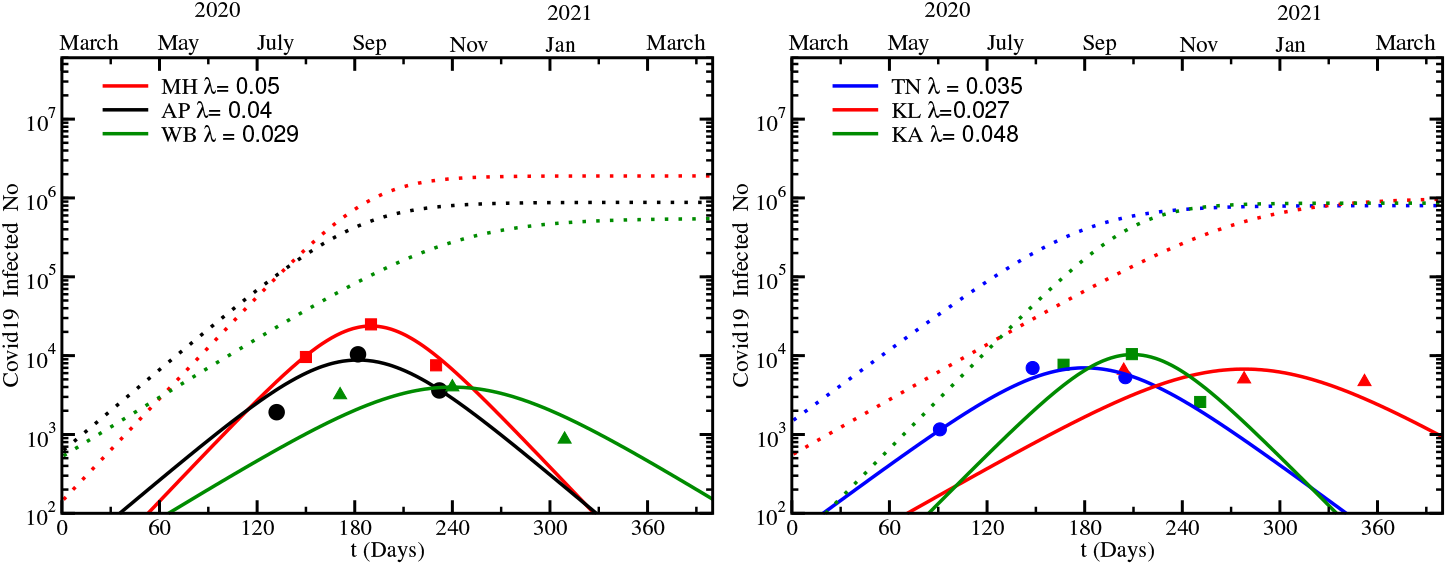
Sigmoid curves(dotted lines) and their derivatives (solid lines) for first wave in MH, AP, WB, TN, KL, KA.

**Figure 3:**
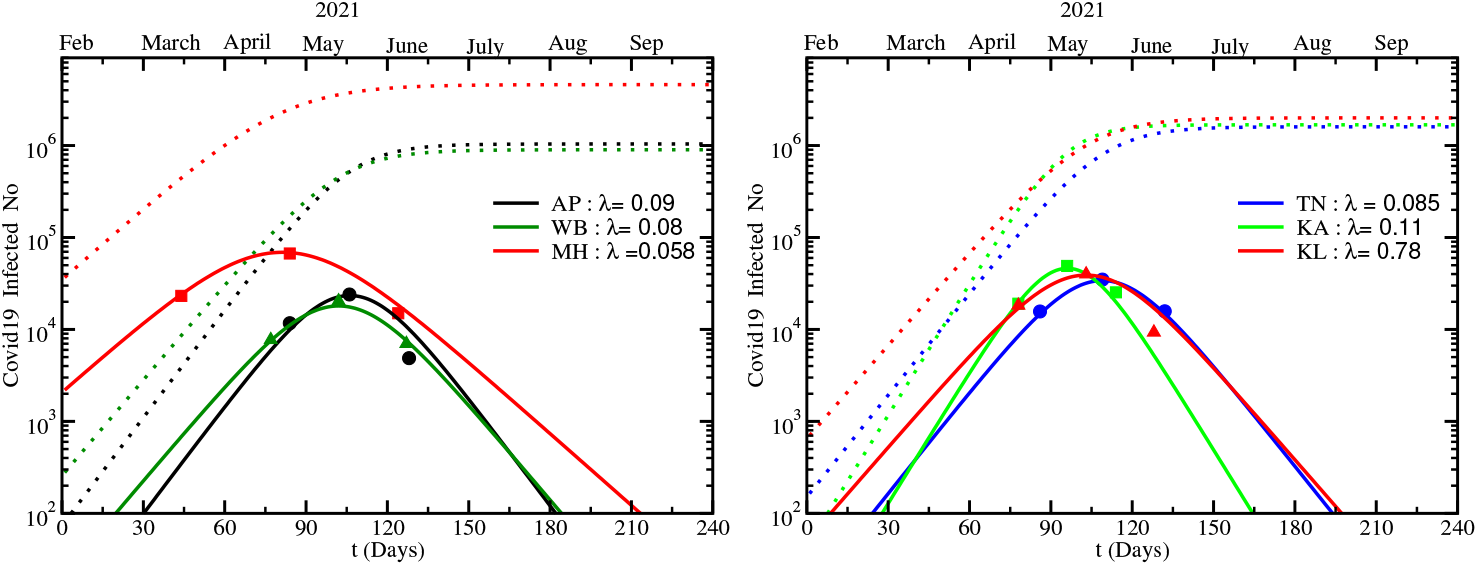
Same as Fig. (2) for second wave.

In first wave, we find the range of growth parameter *λ*_1_ = 0.03 − 0.05 and peak value 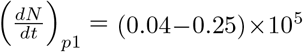. The state level range of growth parameter is quite close to the range of *λ*_1_ = 0.04− 0.05 for entire country. Being added of state level peak values, we find 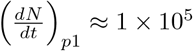 for India. Different states shown peak value at different time points *t*_*p*1_ which are in the range of *t*_*p*1_ = 5 *–* 9 months. India data shows the peak value around *t*_*p*1_ = 6.5 months. If we analyze second wave then state level ranges are *λ*_2_ = 0.078 − 0.1 (excluding MH *λ*_2_ = 0.05), 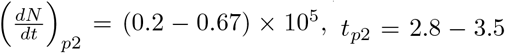 months and India data shows at *λ*_2_ = 0.08, 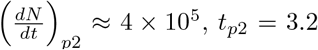 months.

If we compare first and second wave data of India and its different states, then we can notice their ratios as *λ*_2_*/λ*_1_ ≈ 2, *t*_*p*2_*/t*_*p*1_ ≈ 1*/*2, *N*_*m*2_*/N*_*m*1_ ≈ 2. Although ratio between peak values of two waves for different states are not quite stable. As example, it is approximately 4 for India, 5 for WB, TN and 13 for KL etc. Considering India data as collective effect, we may conclude that first to second wave transition was just transition of parameters *λ*_1_ → *λ*_2_ = 2*λ*_1_, *t*_*p*1_ → *t*_*p*2_ = *t*_*p*1_*/*2, *N*_*m*1_ → *N*_*m*2_ = 2*N*_*m*1_ and 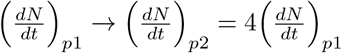.

Now let us come to the question whether we can identify the reason for occurring second wave after the first wave? Single answer of this question is really difficult but mutation of virus might be considered as a dominating point. Here, we will try to understand graphically but reader should considered that quantitative message with a very flexible way. In Fig. (4), we have drawn first and second wave daily cases curves in one portrait and we can see an overlapping/crossing zone of them around February, 2021. This is also observed in data (circles) as one can notice that second wave rising is started after Feb, 2021. We have put few selective daily cases data at 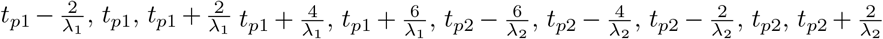. One can notice that our Sigmoid curves for first and second waves are well fitted within 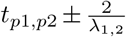. Outside those zones, the trends of data points and curves are going similar although bit of differences in numerical strengths are noticed. Now, assuming mutated virus as a dominating cause of second wave, let us try to describe the data as follows. After entering covid19 virus into India from March, 2020, first wave has received its peak around Sep, 2020 and then it went down until Feb, 2021. This imported covid19 virus are started to be mutated and among the different variant, delta virus [38] become more contagious than previous. We may consider this delta virus as a dominating reason for second wave, which is appeared to be started from Feb, 2021 and received peak around first week of May, 2021. Interestingly, first confirmed case of delta virus in India is observed around Oct, 2020, [39] which is quite earlier than Feb, 2021, from when second wave seems to grow. So it is quite interesting fact that the growing pattern of delta virus was hidden from Oct, 2020 to Feb, 2021. We can only see the decay pattern of first wave curve. If we extend our second wave Sigmoid curve before Feb, 2021, then we get *N*_0_ ≈ 1 around Oct, 2020. Recovering this empirical point by the simple logistic function is really very interesting fact. So if we crudely assign our first and second wave as the outcome of covid19 virus and mutated/delta virus spreading, then we can find recognize their overlapping zone.

**Figure 4:**
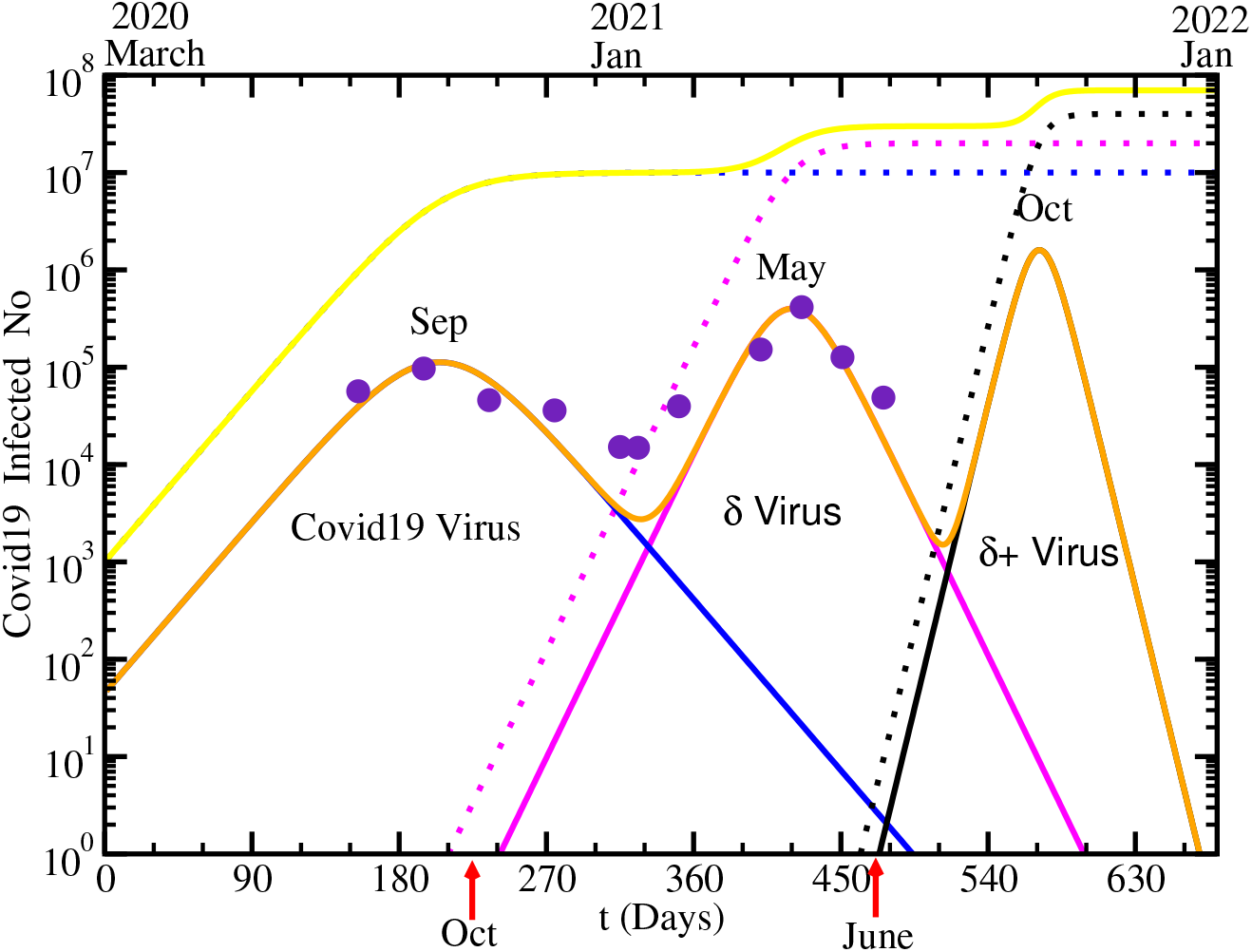
Log scale version of left panel of Fig. (5). Here their superposition are presented by brown and yellow solid lines and starting points of second and third wave are marked by red arrows.

After realizing the transition from first to second wave, we can use the idea for predicting third wave. After mutation of delta virus, recently delta plus (δ+) virus is first reported in India around June, 2021. It is probably 1.5 − 2 times contagious [40, 41] than delta virus, although the fact is not very settled [42]. We already noticed that by simply changing *λ*_1_ = 0.04 and *N*_*m*1_ = 1 × 10^7^ to *λ*_2_ = 0.08 and *N*_*m*2_ = 2 × 10^7^, we get first to second wave transition. Doubling of the growth parameter can be connected with the two times contagious of delta virus with respect to former covid19 virus. So assuming δ+ virus as two times contagious than δ virus and assigning δ+ as a dominating cause for third wave, we have drawn another Sigmoid function with *λ*_3_ = 2*λ*_2_ = 0.16 and *N*_*m*3_ = 2*N*_*m*2_ = 4 × 10^7^ in Fig. (4). We have started it from June with *N*_0_ = 1. As δ virus started to spread from Oct, 2020 but its actual growing become noticeable from Feb, 2021, similarly δ+ virus started in June, 2021 but its actual growing might be found from **mid-Aug, 2021**. Most fearing point is that it might go with rapidly blowing pattern. By transforming log scale to linear scale, Fig. (4) is redrawn in left panel of Fig. (5), from where reader can guess the fearing point. Including our predicted third wave curve along with the observed first and second wave curves, we may present them as transitions among three Sigmoid functions, whose parameter transformation can be expressed in a single equation:

**Figure 5:**
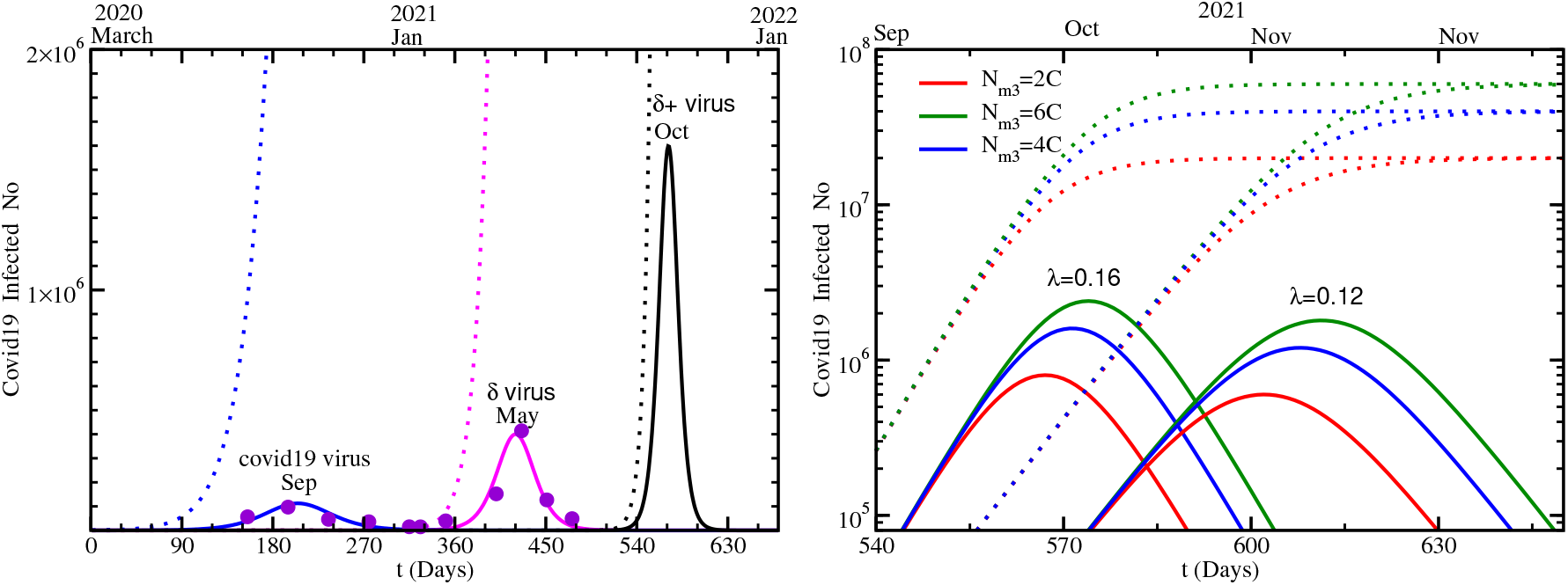
Left: first, second and third wave cumulative (dotted line) and daily (solid line) case Sigmoid curves and few selective data points (circles) of daily cases. Right: Predicted third wave Sigmoid functions - cumulative (dotted line) and daily (solid line) cases for different guess values of growth parameters *λ*_3_ and maximum cumulative value *N*_*m*3_.

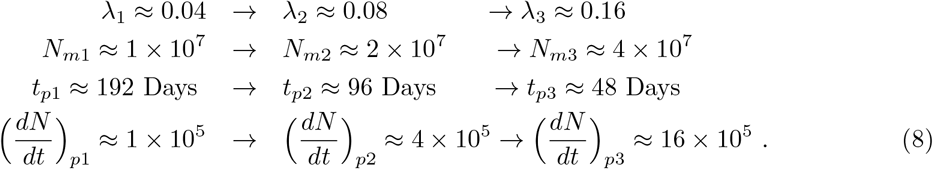

Since there are no empirical data points from where we can guess third wave parameters, so as a possible numerical range, right panel of Fig. (5) is dedicated for that. By taking *N*_*m*3_ = (2, 4, 6)*×*10^7^ and *λ*_3_ = 0.12, 0.16, 6 curves are plotted and we find that the peak values can be varying from 5*×*10^5^ to 23 *×* 10^5^ and peak position can be varying from Sep, 2021 to Nov, 2021.

## 4 Summary

In summary, present work is intended to explain the existing first and second wave covid19 infection data with the help of simple logistic function, called Sigmoid function. From the data points of peak values and peak positions for daily cases of India and its selective states MH, KL, KA, TN, WB, AP, we have found the required input parameters of the Sigmoid functions. Our results grossly indicates a transition between two Sigmoid pattern with twice larger growth parameter and maximum values of cumulative data during first to second wave transition. In parallel, time duration of second wave shrink to half of that first wave and peak values of daily cases becomes four times larger. From the basic properties of Sigmoid functions, those changes can be easily realized.

Another interesting aspect is revealed for mutated delta virus observation. Via backward extensions of second wave Sigmoid curve, we can reach to the initial point (Oct, 2020), when delta virus was first observed. Implementing that idea, we can consider June, 2021 as third wave initial point, since next version mutated delta plus virus is observed on that time. Assuming two times contagious nature delta plus than delta virus, we have considered third wave Sigmoid function with doubled growth parameter with respect to second wave Sigmoid functions. Including our third wave prediction, we have sketched three waves in terms of three Sigmoid functions. Their growth parameters and maximum values of cumulative cases are jumping two times than previous; their daily cases peak values are jumping four time; and their epidemic duration shrinks from 12 months to 6 months to 3 months.

## Data Availability

NA

## Notes

### Competing Interest Statement

The authors have declared no competing interest.

